# Discrimination of bacterial and viral infection using host-RNA signatures integrated in a lab-on-a-chip technology

**DOI:** 10.1101/2022.02.21.22271125

**Authors:** Ivana Pennisi, Ahmad Moniri, Nicholas Miscourides, Luca Miglietta, Nicolas Moser, Dominic Habgood-Coote, Jethro A. Herberg, Michael Levin, Myrsini Kaforou, Jesus Rodriguez-Manzano, Pantelis Georgiou

**Affiliations:** Centre for Bio-Inspired Technology, Department of Electrical and Electronic Engineering, Faculty of Engineering, Imperial College London, UK; Department of Infectious Disease, Faculty of Medicine, Imperial College London, London, UK

**Keywords:** Data-driven Algorithm, ISFET Arrays, Host-response signatures, electronic LAMP, Lab-on-chip, Infectious Disease, Point-of-care diagnostics, Antimicrobial Resistance

## Abstract

The unmet clinical need for accurate point-of-care (POC) diagnostic tests able to discriminate bacterial from viral infection demands a solution that can be used both within healthcare settings and in the field and that can also stem the tide of antimicrobial resistance. Our approach to solve this problem is to combine the use of Host-gene signatures with our Lab-on-a-chip (LoC) technology enabling low-cost LoC expression analysis to detect Infectious Disease.Host-gene expression signatures have been extensively study as a potential tool to be implemented in the diagnosis of infectious disease. On the other hand LoC technologies using Ion-sensitive field-effect transistor (ISFET) arrays, in conjunction with isothermal chemistries, are offering a promising alternative to conventional lab-based nucleic acid amplification instruments, owing to their portable and affordable nature. Currently, the data analysis of ISFET arrays are restricted to established methods by averaging the output of every sensor to give a single time-series. This simple approach makes unrealistic assumptions, leading to insufficient performance for applications that require accurate quantification such as RNA host transcriptomics. In order to reliably quantify host-gene expression on our LoC platform enabling the classification of bacterial and viral infection on chip, we propose a novel data-driven algorithm for extracting time-to-positive values from ISFET arrays. The algorithm proposed is based on modelling sensor drift with adaptive signal processing and clustering sensors based on their behaviour with unsupervised learning methods. Results show that the approach correctly outputs a time-to-positive for all the reactions, with a high correlation to RT-qLAMP (0.85, R2 = 0.98, p < 0.01), resulting in a classification accuracy of 100 % (CI, 95 - 100). By leveraging more advanced data processing methods for ISFET arrays, this work aims to bridge the gap between translating assays from microarray analysis (expensive lab-based discovery method) to ISFET arrays (cheap point-of-care diagnostics) providing benefits on tackling infectious disease outbreak and diagnostic testing in hard-to-reach areas of the world.

## 1. Introduction

Whole blood gene expression biomarkers have been proposed as an accurate and reliable way of patient diagnosis [1]. Scientists worldwide aim to integrate such host-genes as prognostic and predictive marker of disease into clinical diagnostic pathways [2]. Although many signatures have been already described to be linked to infectious disease, their implementation in the clinic as Point-of-care technology is still very limited [3, 4].

In our previous work, we highlighted the potential of a 2-gene RNA signature to be translated into a rapid and portable laboratory-on-a-chip (LoC) tests suitable for Point-of-Care (PoC) applications [5]. In fact, the use of LoCs as diagnostic platforms has been rapidly growing in recent years, especially in resource-limited settings for its potential as reliable, fast, cost-effective and portable solution leading to opportunities in point-of-care application. [6]. In an effort to translate the findings from microarray gene expression discovery to LoC diagnostics, we focused on the detection of RNA from clinical samples and developed a new Machine Learning based algorithm to extract valuable information from an array of electrochemical sensors used as microchip technology to detect the amplification of nucleic acid.

This development is performed with a conventional qPCR instrument using isothermal amplification chemistries, which is then translated on an ion-sensitive field-effect transistor (ISFET) based LoC platform yielding a method referred to as electronic reverse transcriptase quantitative loop mediated isothermal amplification (RT-qLAMP), or RT-eLAMP [5]. Our LoC platform based on metal-oxide-semiconductor (CMOS) technology is indeed in a full development stage and we have already reported the successful detection of different infectious diseases such as *Plasmodium falciparum* cause of Malaria [7], Dengue [8], Zika [9] and most recently SARS-CoV-2 [10].

The vision of this work is two-fold. Firstly, we aim to motivate a workflow from microarray analysis (expensive lab-based discovery) to lab-on-a-chip devices (cheap point-of-care diagnostics), in order to create a more efficient process for RNA transcript detection. Secondly, we show that the vast amount of rich data from ISFET sensor arrays enable a wide variety of algorithms, that can extract as much information as needed in order to provide clinically useful results rather than misleading conclusions. Overall, this work highlights the powerful combination of the host transcriptomic approach combined with our microchip technology to discriminate bacterial from viral infection using a new ML based approach to extract information from ISFET array and provide a diagnostic outcome.

Currently, the data analysis of ISFET arrays is restricted to established methods developed for conventional instruments by averaging the output of every sensor to give a single time-series [11]. All the established methods for detecting amplification and computing a time-to-positive, such as the ‘gold standard’ cycle-threshold method or *C*_*y*0_ method can be used [12, 13]. Moreover, sensor arrays inherently output a spatio-temporal signal and simply averaging sensors assumes that the sensors belong to the same data distribution [14]. This is particularly important when fabricating ISFETs in unmodified CMOS technology due to significant trapped charge effects [15].

In our work, we aimed to classify bacterial and viral infection by exploring a novel data-driven algorithm for extracting time-to-positive values from ISFET arrays (fabricated in unmodified CMOS). We achieved this by modelling sensor drift with adaptive signal processing and clustering sensors based on their behaviour with unsupervised learning methods. First, a 2-gene signature which has been reported to distinguish viral versus bacterial infection using microarray analysis is validated [5]. Subsequently, a novel RT-qLAMP assay is developed for the 2-gene signature based on the splice variants analysis to guarantee a targeted mRNA amplification, high specificity and sensitivity and a lower limit of detection of 10 copies per reaction within 23 minutes. To evaluate real world samples, we have used 24 clinical isolates as a case study (12 with confirmed bacterial infection and 12 with confirmed viral infection), so as to take a step towards tackling antimicrobial resistance.

By leveraging more advanced data processing methods for ISFET arrays, this work hopes to bridge the gap between translating assays from microarray analysis (expensive lab-based discovery method) to ISFET arrays (cheap point-of-care diagnostics). In this work we describe our method for differentiating bacterial from viral infection using RNA host blood expression detected on our lab-on-a-chip technology and applying a novel data-driven algorithm to combat the issues of drift and trapped charge.

## 2. Materials and Methods

### 2.1. LAMP Assay Design

Minimal signatures that discriminate bacterial from viral infection in patients have been used to design LAMP assays to be detected on the LW platform through electronic amplification [5]. Specific LAMP primers for *IFI44L* and *EMR1* were designed to discriminate respectively between viral and bacterial infection. *EMR1* was found over-expressed in patients with bacterial infection, *IFI44L* was found over-expressed in patients with viral infection. To design the LAMP assays six primers were generated to target eight exon-specific region within the gene *IFI44L* and *EMR1*, using Primer Explorer V5 (Eiken Chemical Co. Ltd., Tokio, Japan) Table1. Primers were aligned to the reference genomic sequences using Geneious 10.0.5 software and manually optimised considering their melting temperature (*T*_*m*_), stability at the 3’ end of each primers, GC content, and possible secondary structures such as primer dimer formation using NUPACK37 software (http://www.nupack.org/). The performance of the assays was evaluated based on their analytical sensitivity achieving a lower LoD of 10 copies/reaction. Primers were purchased from Integrated DNA Technologies and resuspended in TE to 100 µM stock solutions. The stocks were stored at −20 °C.

### 2.2. LAMP Reaction Conditions

The optimised pH-LAMP reaction mix comprises: 1 µL of 10 × customized isothermal buffer (pH 8.5 - 9), 0.6 µL of MgSO4 (100mM stock), 0.56 µL of dNTPs (25mM stock), 0.6 µL of BSA (20mg/mL stock), 1.6 µL of Betaine (5 M stock), 0.25 µL of NaOH (0.2 M stock), 0.25 of AMV reverse transcriptase by Promega (25,000 U/mL stock), 0.10 µL of RNAse inhibitor (20,000 U/mL stock), 0.25 µL of SYTO 9 Green (20 µM stock), 0.042 µL of Bst 2.0 DNA Polymerase (120,000 U/mL stock), 1 µL of 10ù LAMP primer mixture, 1 µL of extracted RNA and enough nuclease free water (Thermo Fisher Scientific) when necessary, to bring the volume to 10 µL reactions. All reagents except the AMV Reverse Transcriptase were purchased from New England BioLabs. Reactions were performed at a single temperature of 63 °C for 30 min in the LW platform and validated using the same reaction condition into LightCycler 96 (Roche Diagnostics) by using a LightCycler 96 (LC96) Real-Time PCR System (Roche Diagnostics). Furthermore, validation of the specificity of the products was performed running one melting cycle in the LC96 at 0.1 °C_s from 65 °C up to 97 °C for validation of the specificity of the products. Each experimental condition was run in triplicates. For RT-eLAMP, the reagents were modified to obtain a low buffered solution with the aim to detect nucleotides by measuring the solution’s pH change. The reaction mixture contains the same reagents except the Tris buffer, which was removed to obtain a low buffered solution, where pH change can be measured. To carry-out DNA amplification reactions on chip the microfluidic chamber was filled with 10 µL of the same reaction mix prepared for the gold standard instrument and sealed with PCR tape to avoid evaporation and contamination of the amplified products while running on the platform.

### 2.3. Conventional method

The conventional method of analysing amplification data from ISFET arrays typically revolves around averaging the signals from the sensors and subsequently processing the data (e.g. drift compensation). Therefore, herein, the conventional method is referred to as *average then process*, or ATP. The ATP method, described in Malpartida et al [7], is shown below and the key assumptions which the proposed method aims to tackle are boxed in gray.

**Table 1.**
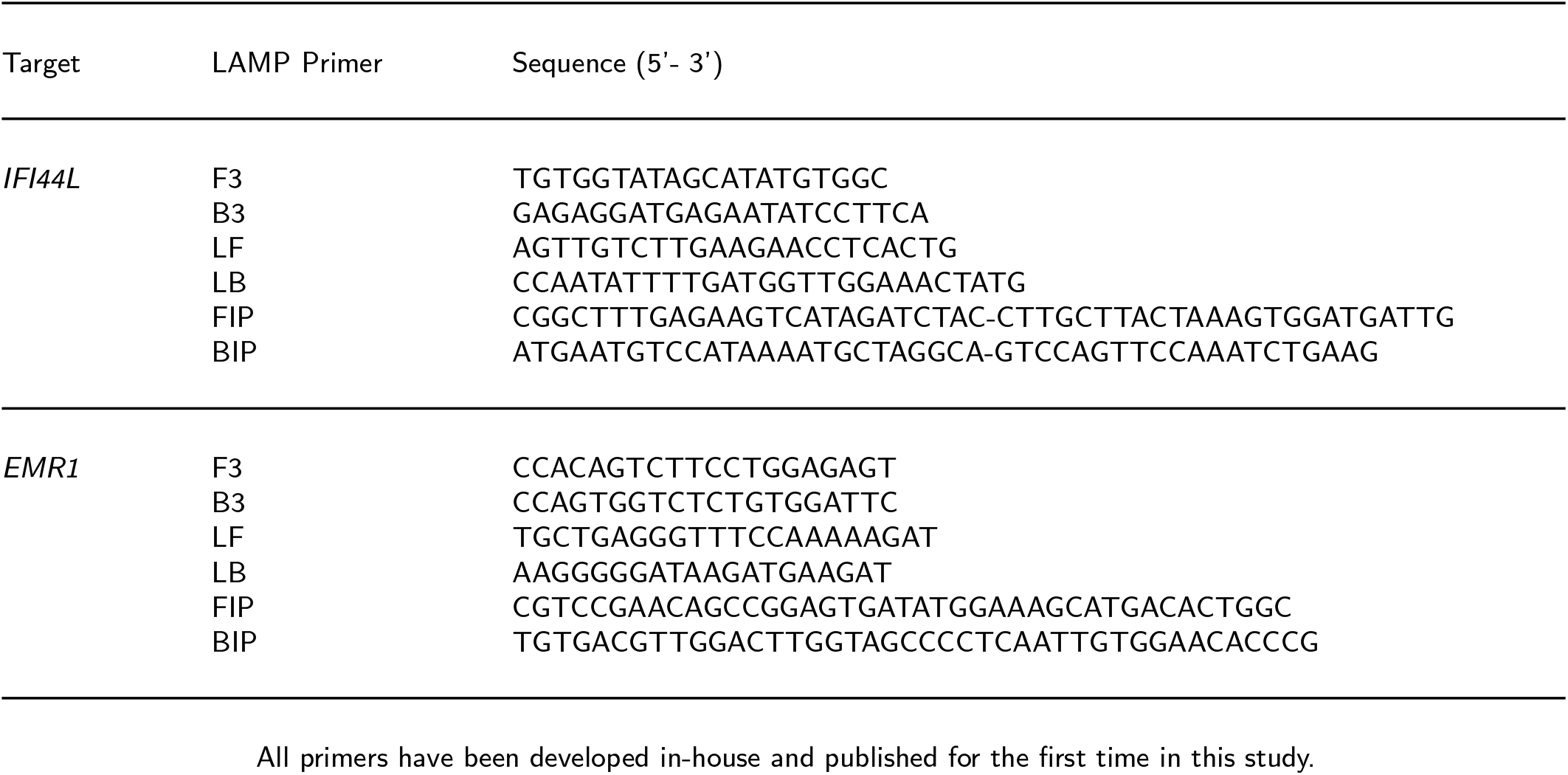
LAMP Primer sequences for *IFI44L* and *EMR1* targets.

1. The signals from all ISFETs in contact with the chemical solution are averaged. The resulting signal is normalised by removing the DC component.

#### Assumption 1

The signal from all the sensors are drawn from the same distribution.
2. During the first few minutes of the chemical reaction, sensor drift is modelled using a stretched exponential function described by:

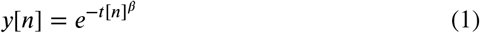

Where *y*[*n*] and *t*[*n*] are the voltage and time values at sample *n*, and *β* is a scalar parameter. The drift is extrapolated across time, and subsequently subtracted from the signal to yield an amplification curve.

#### Assumption 2

The drift parameter *β* is stationary, or equivalently, *β* does not change with time.
3. The time-to-positive is extracted as described in Guescini et al (2008) [16] via the *C*_*y*0_ method.

### 2.4. Proposed Method

The proposed method, as depicted in Figure 2, follows 5 steps, namely: (i) pre-processing, (ii) adaptive signal processing, (iii) unsupervised clustering, (iv) spatial validation and (v) averaging & peak detection. Each step is described below.

**Figure 1:**
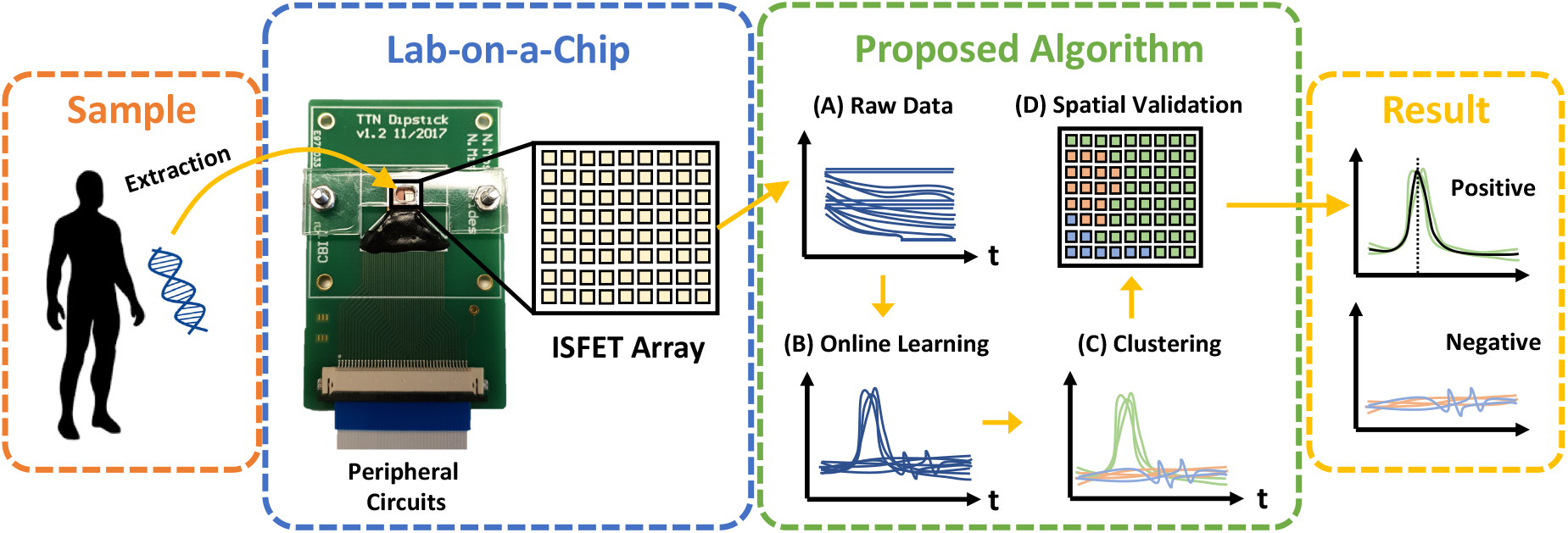
Depiction of the sample-to-result workflow using a lab-on-a-chip device in conjunction with the proposed algorithm

**Figure 2:**
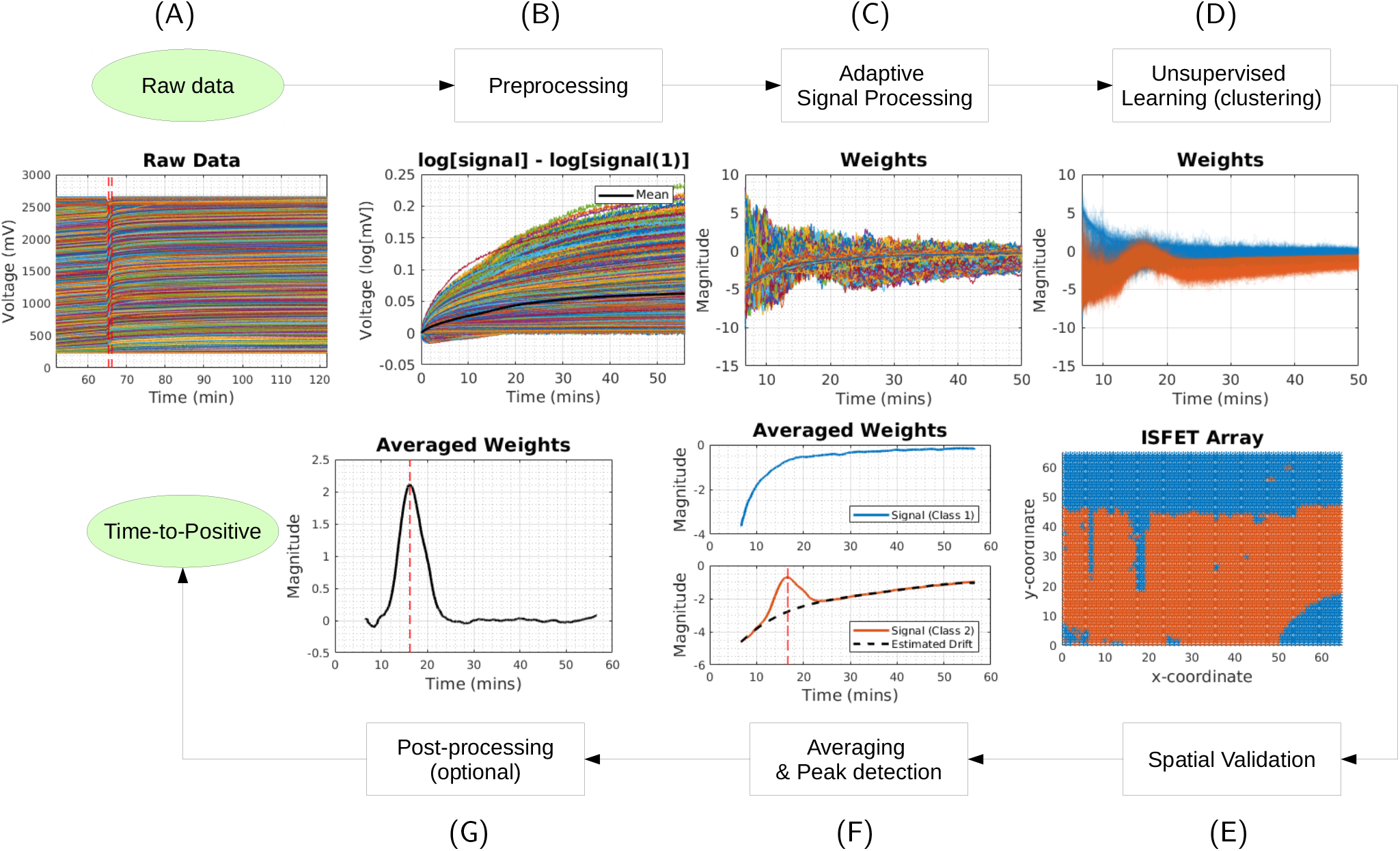
Proposed method for extracting time-to-positive values from ISFET arrays

**Pre-processing**. The background is removed from each signal by subtracting the average of the first 5 voltage measurements. An example of raw data and pre-processed signals are shown in Figure 2 (A) and (B). **Adaptive Signal Processing**. The drift is typically modelled via the stretched exponential given in equation 1. The parameter *β*is estimated via any iterative gradient based optimisation algorithm, commonly of the form:

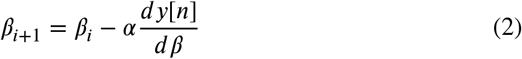

Where *i* is the iteration number, α is a small constant (note: this can be adaptive), and 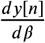 can be computed as:

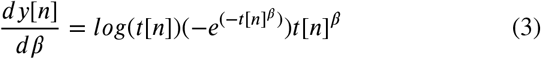

The above analysis assumes that the drift parameter is constant through time. In order to relax this assumption, the proposed method takes an ‘adaptive signal processing’ approach whereby the drift parameter is updated at each time step, so as to capture the behaviour over time. Therefore, each sensor will yield a *β*vector with the same length as the experiment, as depicted in Figure 2 (C). In this study, the normalised least mean square is used [17]. **Unsupervised Clustering**. The *β*vectors are grouped into *K* clusters via K-Means clustering using the cosine distance, whereby the value *K* is determined via the silhouette score [18]. Therefore, sensors are grouped by similar behaviour, as depicted in Figure 2 (D). **Spatial Validation**. In reality, we expect sensors which are spatially close to have more similar behaviour. Therefore, to ensure the clustering is not random, the clusters can be visualised spatially, as in Figure 2 (E), to confirm this. This step can be achieved automatically via graph theoretic approaches for calculating the number of components [18]. **Averaging & Peak Detection**. The signals in each cluster are averaged to remove the effect of inherent noise, and a peak detection algorithm is used to extract a time-to-positive value, as shown in Figure 2 (E). Note: Since the behaviour of sensors are commonly drifting over time, an optional post-processing is to remove slow components in the signals, as in Figure 2 (F).

For reproducibility purposes, the RNA targets, primer design, reaction conditions and statistical analysis are described below. Details of the microarray analysis can be found in Herberg et al. (2016) [1] and the ISFET experimental setup can be found in Malpartida et al. (2019) [7]. The reader may wish to skip directly to the Results and Discussion (section 3).

### 2.5. Microarray analysis

Values for gene expression data analyzed on Illumina microarrays were provided by Herberg and colleagues [1]. Using the data provided, we identified the clinical samples that had differentially expressed transcripts between the definite viral and bacterial groups with |log2 FC| > 1 and adjusted P-value < 0·05, using a trained support vector machine classifier. These thresholds were chosen to ensure that differential expression for selected variables could be distinguished using the resolution of other validation techniques (i.e. qPCR) TBC

### 2.6. Statistical Analysis

To define a decision boundary between the bacterial and viral infections, a support vector classifier from the scikitlearn package in Python was used [19]. The underlying regression, i.e. linear combination of time-to-positive values were used as a score. To prevent over-fitting, 10-fold cross-validation was used to select hyper-parameters. The confidence interval for the classification accuracy was determined by 95% under the binomial distribution. The correlation between methods was determined using the Pearson correlation and a p-value was determined using the F-statistic.

## 3. Results and Discussion

The following section is structured as follows. First, the 2-gene signature for distinguishing bacterial and viral infections is validated using microarray analysis. Subsequently, a new RT-qLAMP assay for detecting the same 2-gene is demonstrated. Following this, the ATP and proposed algorithm for extracting time-to-positive values from ISFET arrays are evaluated. Finally, the correlation between moving from microarray analysis to RT-qLAMP, and from RT-qLAMP to RT-eLAMP is studied to highlight the bottlenecks in the translation process.

### 3.1. Microarray Analysis

Figure 3 (A) shows the 2-gene signature (*IFI44L* and *EMR1*) for 11 bacterial and 11 viral isolates. It can be observed that the two groups are linearly separable. In particular, Figure 3 (A) shows the decision boundary for a trained support vector machine classifier with a black dashed line. The classification accuracy, as determined by cross-validation, is shown to be 100.0% (CI, 95.7 − 100.0%). Figure 3 (B) shows boxplots with scores which corresponding to the euclidean distance between each data point and the decision boundary. The log RNA expression can be found in Figure 3 (C). This analysis demonstrates that the 2-gene signature is in fact a successful method of distinguishing viral versus bacterial infection in microarray analysis. However, detection with gene expression microarrays needs sophisticated lab equipment, is expensive and cannot be used for routine diagnostics.

**Figure 3:**
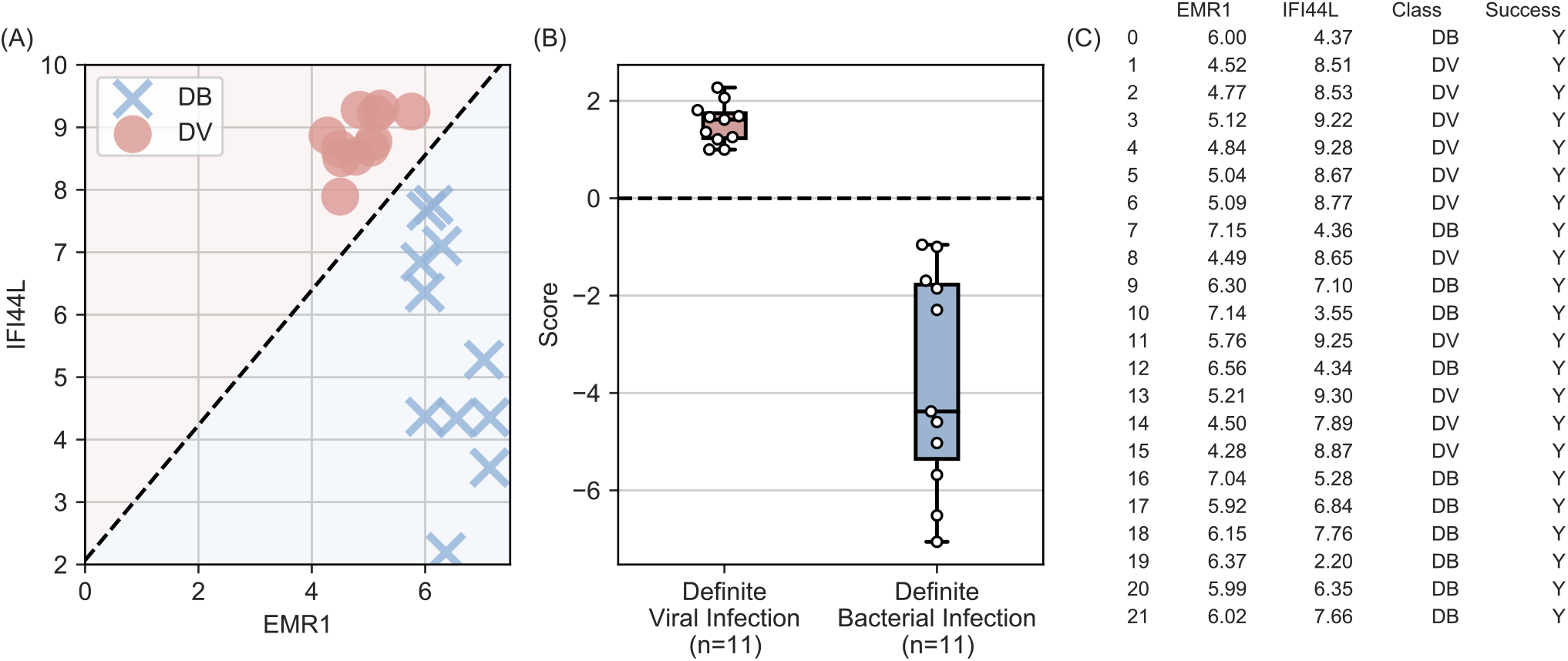
Viral versus bacterial classification - performance of microarray analysis

### 3.2. RT-qLAMP

To this end, two new LAMP assays were developed to detect the 2-gene signature using a conventional qPCR platform. Figure 4 shows the standard curves for *IFI44L* and *EMR1* for concentrations ranging from 10 to 10^6^ copies per reaction, respectively. This demonstrates that the assay is highly sensitive with a lower limit of detection down to 10 copies per total volume reaction. With regards to classification performance, Figure 5 illustrates that RT-qLAMP assay also provides a 100.0% (CI, 95.7−100.0%) accuracy, showing the successful translation across platforms. Since microarray measures the RNA abundance, it is expected that the time-to-positive value from LAMP is inversely proportional (i.e. high expression results in faster reaction). As a sanity check, it can be observed that the position of DB and DV isolates are swapped in 2D space (i.e. for microarray, DB is in the lower right and DV is in the upper left, and vice-versa for RT-qLAMP).

**Figure 4:**
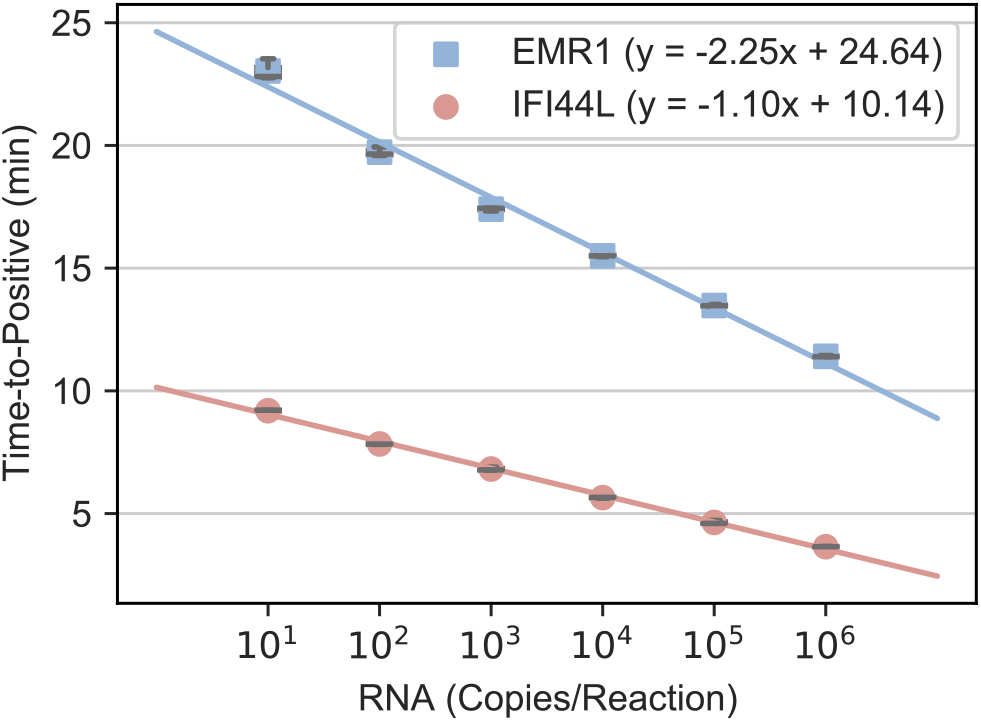
RT-qLAMP standard curves for *IFI44L* and *EMR1*

**Figure 5:**
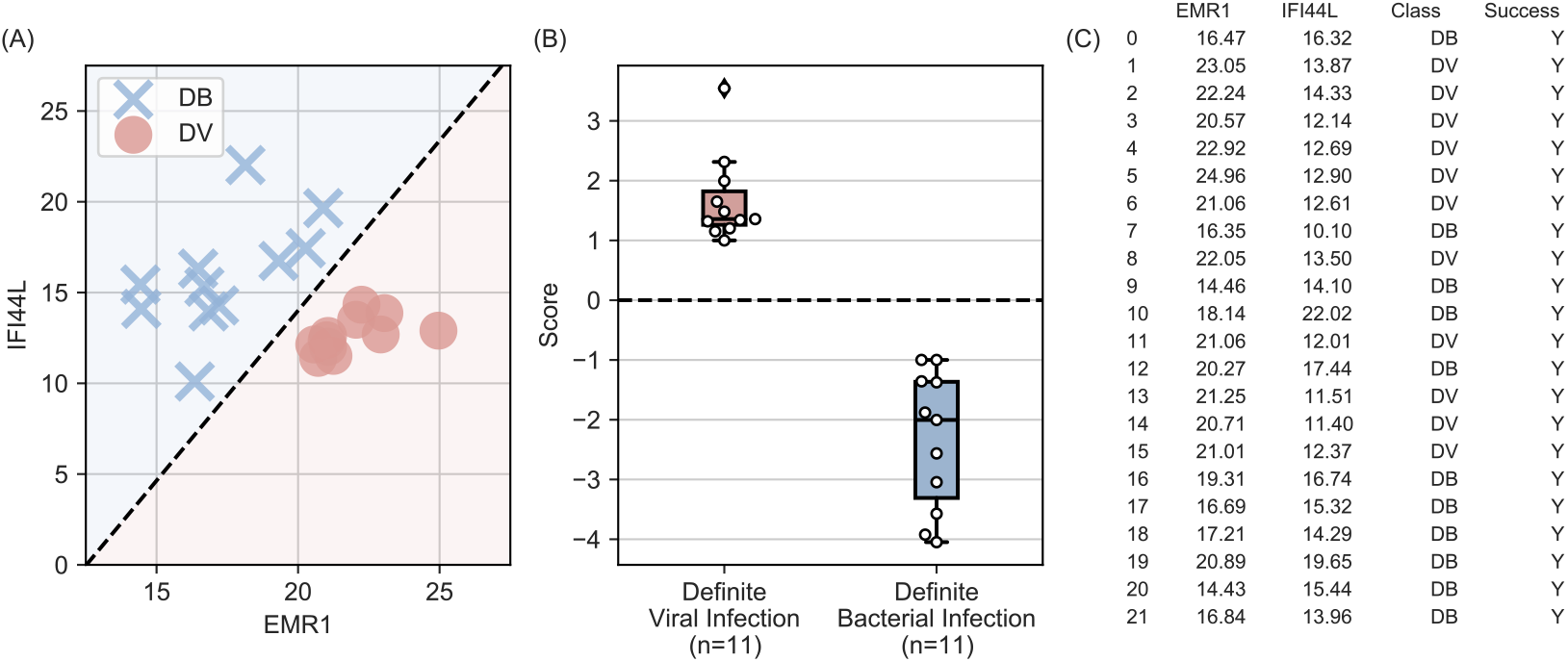
Viral versus bacterial classification - performance of RT-qLAMP analysis

### 3.3. Electronic RT-LAMP

Subsequently, the same assays and clinical isolates were tested on the ISFET-based LoC platform using the ATP algorithm. The results are shown in Figure 6. It can be observed from 6 (C) that 36.4% of the experiments were considered negative/failed, resulting in 50% of the isolates failing to provide a 2D signature. This highlights the challenge presented with ISFETs which have significant noise. Moreover, it can be observed from Figure 6 (A) that due to the lack of data, the decision boundary has a negative slope coefficient, which is not correct. It is important to stress that this method is sufficient where most of the sensors have similar behaviour (e.g. low trapped charge and noise effects), as in Malpartida et al [7]. In contrast with the ATP method, the proposed method successfully called positive for all experiments. As shown in Figure 7, the classification accuracy is 100.0% (CI, 95.7 − 100.0%) with a positive decision boundary slope, although the class separation is not as evident as microarray analysis or RT-qLAMP.

**Figure 6:**
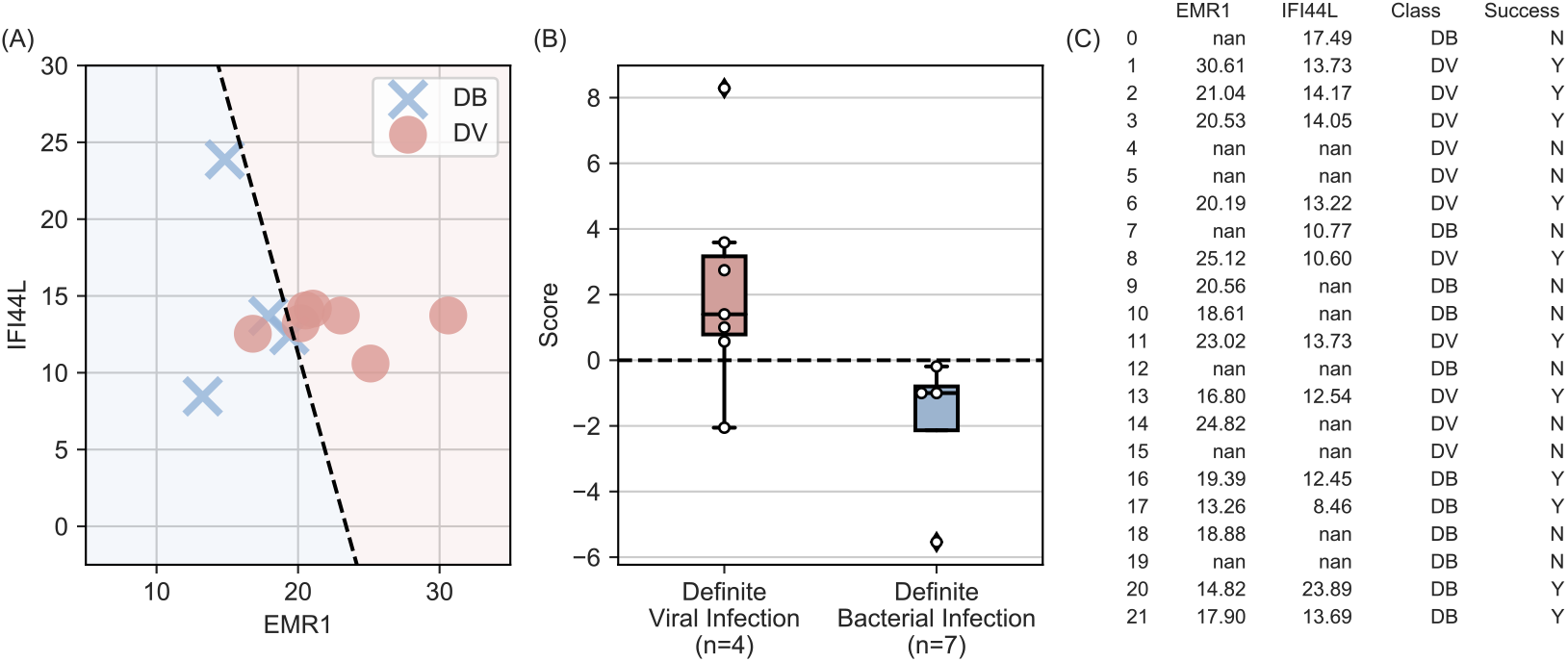
Viral versus bacterial classification - performance of RT-eLAMP analysis (ATP method)

**Figure 7:**
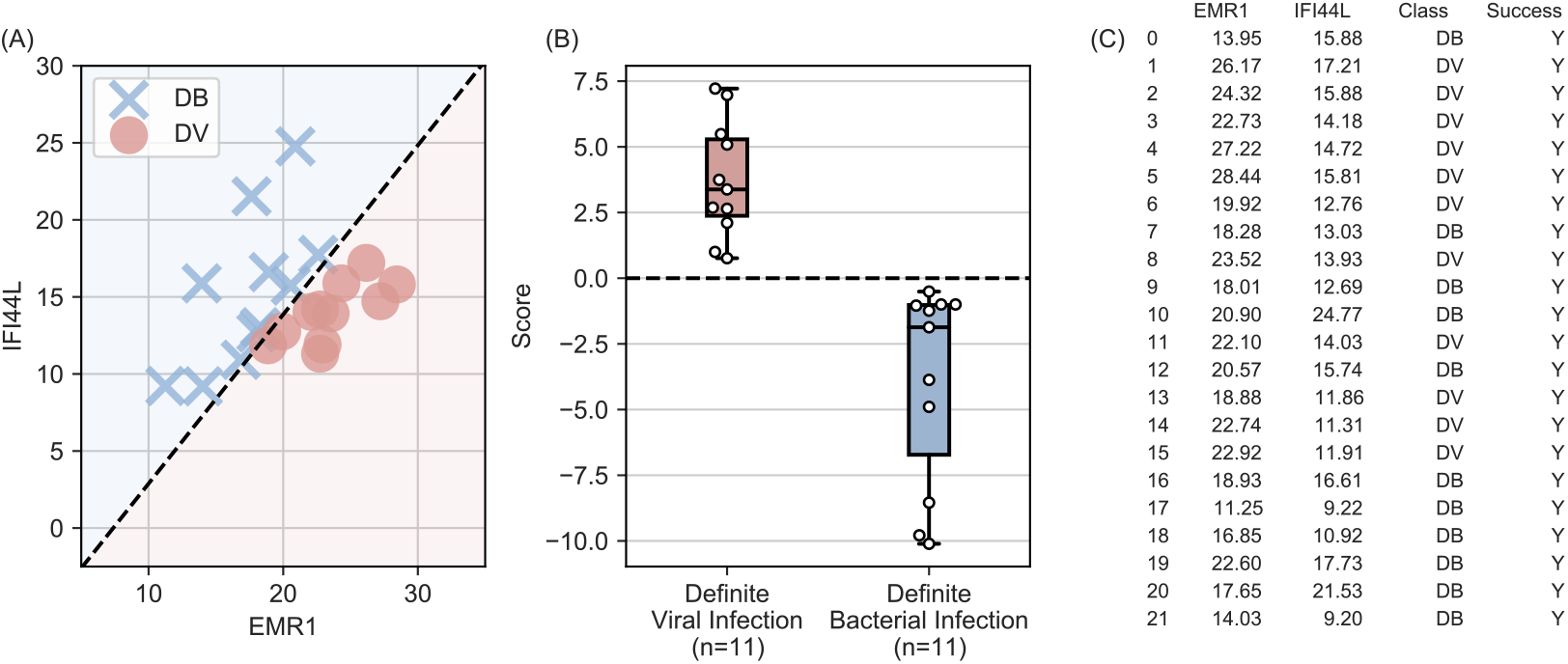
Viral versus bacterial classification - performance of RT-eLAMP analysis (proposed method)

### 3.4. Evaluating the Translation Process

Although it has been shown that RT-qLAMP and RT-eLAMP can be used as a viable diagnostic solution for this specific application, microarray analysis remains a useful tool for discovering the signatures. Therefore, it is important to study the translation process from microarray analysis to RT-qLAMP to RT-eLAMP. This is particularly useful for identifying the bottlenecks in the process, so as to improve the class separation. Figure 8 shows a scatter plot between the RNA expression from microarray analysis and RT-qLAMP Cq values, and a scatter plot between RT-qLAMP and RT-eLAMP Cq values. The correlation coefficient between microarray analysis and RT-qLAMP is found to be −0.71 with *R*^2^ = 0.51 and *p*-value << 0.01. However, the correlation between RT-qLAMP and RT-eLAMP is 0.85 with *R*^2^ = 0.98 and *p*-value << 0.01. This demonstrates that the bottleneck in the translation process is between microarray analysis and qPCR, highlighting the need to focus research efforts on molecular assay development. On the other hand, the proposed algorithm correctly outputs a time-to-positive for all the reactions, with a high correlation to RT-qLAMP (0.85, R^2^ = 0:98, p < 0:01), and resulting in a classification accuracy of 100 (CI, 95 − 100).

**Figure 8:**
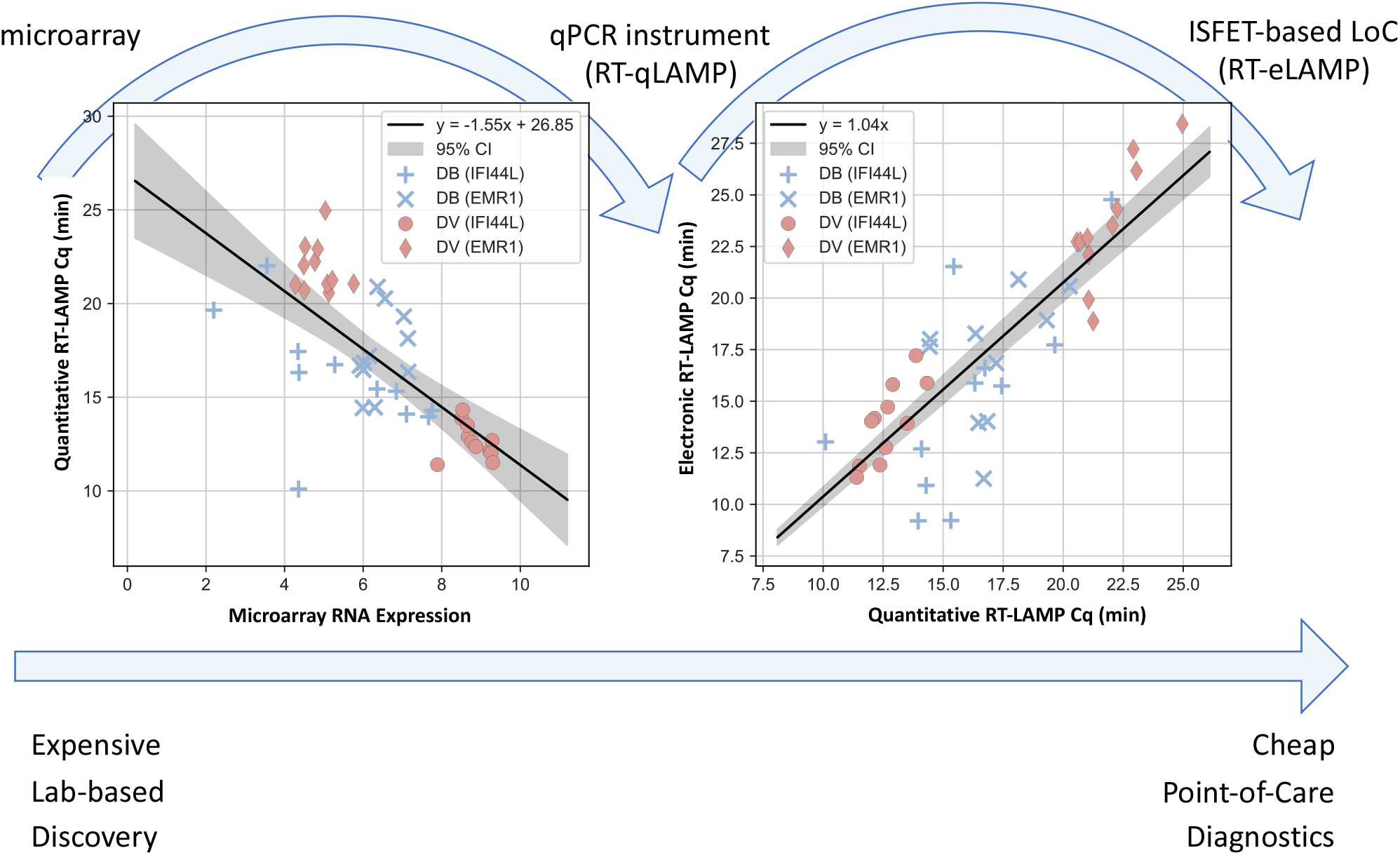
Translation of 2-Gene Signature from microarray analysis to RT-qLAMP to RT-eLAMP - moving from expensive lab-based discovery to cheap point-of-care diagnostics

## 4. Conclusion

There is an urgent need for fast, cheap, robust and quantitative methods for distinguishing between different types of infectious or inflammatory diseases [1]. When they cannot be diagnosed in a timely manner, subjects may receive unnecessary treatments or risk misdiagnosis [20]. Among the main classes of diagnostic methods utilised for detection and identification of infectious diseases either relying on pathogen or host-based diagnosis, only nucleic acid-based methods have a quantitative rather that qualitative output. However, these techniques rely on lab-based instruments such as microarrays, RNA sequencing and qPCR and are also expensive and cannot be miniaturized into a small form factor device requiring a technically trained operator [21]. Furthermore, as sensing systems become more advanced, the extracted data is becoming richer and more complex, presenting new challenges. In the case of diagnostic systems based on ISFET arrays, a major challenge is to combat the effects of various sources of noise [22]. In this study we addressed these and other disadvantages by providing an affordable solution for quantitative detection of RNA-host transcripts highly expressed in the presence of specific diseases. The label-free detection method based on scalable microchip semiconductor technology combined with isothermal amplification chemistries makes this approach suitable to implementation at low-cost. A new data-driven system to identify a nucleic acid amplification can significantly improve the robustness compared with existing methods and the whole system can be delivered at the point-of-care level without the need for large and expensive laboratory equipment, while still delivering a quantitative result that can lead to a diagnostic outcome.

Although this study is a first step towards exploring new methods of implementing gene expression signature into a Point-of-care application and extracting time-to-positive values from ISFET arrays to provide more accurate outcome, there are several factors which should be highlighted for future studies and improvements. In fact, it is important to understand the cost of using the proposed algorithm from computational and memory complexity. In terms of computational complexity, the added operations include: (i) fitting a drift model to each sensor, as opposed to the average of all sensors; (ii) unsupervised clustering such as the Kmeans algorithm; (iii) spatial validation; and (iv) peak detection. For (i), the operations consist of simple additions and multiplications, however the complexity scales with an increase in the number of sensors might result expensive for large arrays. On the other hand, the computation for each sensor is independent and therefore can be completely parallelised, yielding the same complexity as conventional drift fitting. The complexity of (ii)-(iv) largely depends on the chosen algorithms. Although with modern computers, these are not crucial. For example, entire proposed algorithm (without visualisation) took only 0:96, 0:11 seconds on a MacBook Pro (16-inch, 2019, 2.3 GHz 8-Core Intel Core i9 processor and 32 GB 2667 MHz DDR4 memory) running Matlab R2019B. In terms of space complexity, the additional memory usage is in the order of 10 Mb. Aside from algorithmic factors, it is also important to evaluate the experimental design of the study. This includes the experimental conditions and sample set. Regarding the conditions, the experiments were performed similar to a real setting with the exception of using a thermocycler for thermal management. This was in fact a study design choice in order to remove noise from temperature effects since this can be fixed using good thermal control, rather than increasing algorithmic complexity by accounting for temperature. However, a full study including this source of noise should be included in future studies. Moreover, this study used 24 clinical isolates (12 bacterial and 12 viral), and although confidence intervals were used to estimate the upper and lower bounds of the classification accuracy, a larger clinical study is required to ensure there was no unintended sampling bias.

In conclusion, a new method for extracting time-to positive values from nucleic acid amplification for ISFET arrays under high noise environments is proposed. This approach is primarily based on modelling the sensors adaptively to represent ‘changes in behaviour’, and clustering the time-series, so as to average sensors with the same behaviour. Using this new method was shown to successfully provide a high correlation between conventional lab-based RT-qLAMP and the ISFET-based LoC platform, yielding 100.0% accuracy when classifying viral and bacterial isolates with a 2-gene signature. Moreover, the entire workflow from microarray analysis (expensive lab-based discovery) to lab-on-a-chip devices (cheap point-of-care diagnostics) was studied, in order to create a more efficient process for nucleic acid detection. With our work we want to encourage researchers to apply this methodology to other applications, such that it benefits the wider scientific community. We indeed strongly believe that our approach will enable prompt, optimised care which minimises the use of unnecessary treatments such as antibiotics, and instead guides rapid decisions on the use of antiviral or adjunctive treatments including anti-inflammatory drugs with the advantage for surveillance of infection. Furthermore, accurate diagnosis could inform real-time bed planning and escalation of care pathways and ventilator access/allocation improving safety for patients and for healthcare staff. And lastly establishing the utility of an affordable and accurate device that is based on host-transcript signatures will address many diagnostic and prognostic clinical challenges for various infections and inflammatory conditions and others pathology for which there is a blood diagnostic signature. This will have impact on other infectious disease outbreaks, and on diagnostic testing in hard-to-reach areas of the world.

Future directions of this study will focus on the embedded integration of the proposed algorithm into the PoC device itself (rather than running on a computer or server) and a systematic pipeline for *in silico* development of LAMP assays from microarray experiments.

## Data Availability

All data produced in the present work are contained in the manuscript

## Acknowledgements

This work has been funded by Rosetrees Trust (grants P78740 and P78699). Mr Moniri is funded by the Engineering and Physical Sciences Research Council Doctoral Training Partnership (EP/N509486/1). Dr Kaforou is funded by the Wellcome Trust (Sir Henry Wellcome Fellowship grant 206508/Z/17/Z). Drs Kaforou, Herberg, and Levin receive support from the National Institute for Health Research Imperial College Biomedical Research Centre (grant DMPED P26077).

## CRediT authorship contribution statement

**Ivana Pennisi:** Concept and design, Acquisition, analysis, or interpretation of data, Drafting of the manuscript. **Ahmad Moniri:** Concept and design, Acquisition, analysis, or interpretation of data, Drafting of the manuscript, Statistical analysis. **Nicholas Miscourides:** Concept and design, Acquisition, analysis, or interpretation of data. **Luca Miglietta:** Acquisition, analysis, or interpretation of data, Drafting of the manuscript. **Nicolas Moser:** Acquisition, analysis, or interpretation of data. **Dominic Habgood-Coote:** Concept and design, Acquisition, analysis, or interpretation of data, Statistical analysis. **Jethro A. Herberg:** Acquisition, analysis, or interpretation of data, Obtained funding. **Michael Levin:** access to all of the data in the study and take responsibility for the integrity of the data and the accuracy of the data analysis. **Myrsini Kaforou:** Concept and design, Acquisition, analysis, or interpretation of data, Statistical analysis, Obtained funding. **Jesus Rodriguez-Manzano:** Concept and design, Drafting of the manuscript, Obtained funding. **Pantelis Georgiou:** Concept and design, access to all of the data in the study and take responsibility for the integrity of the data and the accuracy of the data analysis, Obtained funding.

## Notes

### Competing Interest Statement

The authors have declared no competing interest.

### Author Declarations

The source data are openly available and referred in the manuscript: JAMA. 2016;316(8):835-845. doi:10.1001/jama.2016.11236

